# Self assessment overestimates historical COVID-19 disease relative to sensitive serological assays: cross sectional study in UK key workers

**DOI:** 10.1101/2020.08.19.20178186

**Authors:** Ranya Mulchandani, Sian Taylor-Philips, Hayley E Jones, AE Ades, Ray Borrow, Ezra Linley, Peter D Kirwan, Richard Stewart, Philippa Moore, John Boyes, Anil Hormis, Neil Todd, Antoanela Colda, Ian Reckless, Tim Brooks, Andre Charlett, Matthew Hickman, Isabel Oliver, David Wyllie

## Abstract

**Objective:** To measure the association between self-reported signs and symptoms and SARS-CoV-2 seropositivity.

**Design:** Cross-sectional study of three key worker groups.

**Setting:** Six acute NHS hospitals and two Police and Fire and Rescue sites in England.

**Participants:** Individuals were recruited from three streams: (A) Police and Fire and Rescue services (n = 1147), (B) healthcare workers (n = 1546) and (C) healthcare workers with previously positive virus detection (n = 154).

**Main outcome measures:** Detection of anti-SARS-CoV-2 antibodies in plasma.

**Results:** 943 of the 2847 participants (33%) reported belief they had had COVID-19, having experienced compatible symptoms (including 152 from Stream C). Among individuals reporting COVID-19 compatible symptoms, 466 (49%) were seronegative on both Nucleoprotein (Roche) and Spike-protein (EUROIMMUN) antibody assays. However, among the 268 individuals with prior positive SARS-CoV-2 tests, of whom 96% reported symptoms with onset a median of 63 days (IQR 52 – 75 days) prior to venesection, Roche and EUROIMMUN assays had 96.6% (95% CI 93.7% – 98.2%) and 93.3% (95% CI 89.6% – 95.7%) sensitivity respectively. Symptomatic but seronegative individuals had significantly earlier symptom onset dates than the symptomatic seropositive individuals, shorter illness duration and a much lower anosmia reporting frequency.

**Conclusions:** Self-reported belief of COVID-19 was common among our frontline worker cohort. About half of these individuals were seronegative, despite a high sensitivity of serology in this cohort, at least in individuals with previous positive PCR results. This is compatible with non-COVID-19 respiratory disease during the COVID-19 outbreak having been commonly mistaken for COVID-19 within the key worker cohort studied.

**What is already known on this topic:** Screening for SARS-CoV-2 antibodies is under way in some key worker groups; however, how this adds to self-reported COVID-19 illness is unclear. There are limited studies that investigate the association between self-reported belief of COVID-19 illness and seropositivity.

**What this study adds:** About one third of a large cohort of key frontline workers believed they had had COVID-19 infection. In around half of these there was no serological evidence of infection. Individuals who believed they had previous infection, but were seronegative, differed systematically from the seropositive individuals: disordered sense of taste and smell was less common, illness duration was shorter, and reported onset of illness commonly predated the main COVID-19 epidemic in the UK.

Although some individuals with previous COVID-19 may be seronegative, among symptomatic individuals who had PCR-confirmed SARS-CoV-2 within our cohort, sensitivity of the two immunoassays used (Roche Elecsys ® and EUROIMMUN) exceeded 90%. Together, these data indicate that many key workers may falsely believe, based on symptomatic illness experienced during 2020, that they have had COVID-19. Further research investigating the relationship between antibody detection and protection from future infection, with and without a history of COVID-19 disease, will help define the role serological testing can play in clinical practice.

## Introduction

After SARS-CoV-2 infection, most individuals mount antibody responses^1 2 3 4 5,6^. Serological tests aim to identify people who have previously been infected, through detection of anti-SARS-CoV-2 antibodies^7^. Currently, serological tests are important in helping us better understand how the disease has spread in the population, and can support pandemic planning and response^8^. In the future they may also be used for individual risk assessment; however at present, although antibody titre correlate with *in vitro* neutralisation^1,3^ and clinical studies suggest an antibody-protection association^9^, it has not yet been proven whether presence of antibody indicates protection against future infection; as such, the usefulness of serology testing in clinical practice is currently unclear.

Early in the response to the epidemic, PCR testing was restricted to those needing hospital care. As testing provision rose, test availability was progressively extended to include health care workers, and (less extensively) to other key worker groups such as Police officers. Only in the latter stages of the first wave of infection was widespread community testing available to all who developed symptoms; initially this was restricted only to those with fever or cough. Collectively, this may have resulted in parts of the population believing they have had COVID-19 because of illness during the pandemic, some of whom may not have had the virus. However, the validity of this understandable assumption, and whether it is justified given particular symptom combinations, is poorly quantified. Although some COVID-19 symptoms, such as taste/smell disorders, are highly specific for COVID-19^10^, many others occur in other viral respiratory infections, and a recent Cochrane review has highlighted the substantial uncertainty about the value of clinical symptoms in the diagnosis of COVID-19^11^. Several studies have looked at the association between individual symptoms and seropositivity^12,13^, identifying associations with symptoms such as anosmia and ageusia and seropositivity; however, to our knowledge, no study has assessed the relationship between self-reported belief of previous COVID-19, combinations of signs and symptoms, and seropositivity.

The UK Government recently launched a mass antibody testing programme for staff in the National Health Service (NHS) and for care workers^14^. Large scale testing on historical sera has indicated a very high specificity of the assays used^15^, but concerns have been raised about a lack of data on assay performance > 35 days post infection^7^, in community cases (particularly those that did not meet testing criteria early in the response), and serological responses which may be of short duration^16^. Here we address existing uncertainties as to value of symptoms^11^ in diagnosing former COVID-19, comparing symptom based self-diagnosis with that of serological detection of symptomatic illness more than 35 days post infection, and quantify the performance.

## Methods

### Study population

The study population consists of various frontline key workers, who are anticipated to be the initial users of UK government home antibody testing programme. Key workers are targeted for recruitment as part of the EDSAB-HOME study, a programme designed to evaluate the accuracy of point of care antibody tests. We recruited three streams of frontline workers with differing expected seroprevalence, severity and ongoing risk of exposure: (A) Police and Fire – many of whom are public facing, (B) HCW – many of whom are patient and/or client facing, and; (C) HCW-PP who had a previous positive nasal or throat swab for SARS-CoV-2 (with or without symptoms) and therefore are confirmed to have had previous COVID-19. In this paper, our focus is on the relationship between signs and symptoms and seropositivity. RT-PCR testing was undertaken as part of the UK COVID-19 testing programme, not as part of the study, and results were self-reported by participants.

### Recruitment and data collection

Prospective workplace based recruitment was conducted between 01 and 26 June 2020. Senior staff in the NHS, Police and Fire organisations were invited to a telephone conversation in which the study was described and invited to support the study. Those who agreed sent out an ethically approved advert by email to all staff. Further methodological details are in Supplementary Material.

Individuals interested in the study were directed to an online study portal, which included the participant information sheet. If they agreed to take part, they completed an online consent form and epidemiological questionnaire (Supplementary Material), regarding their previous exposures, whether they believed they had had COVID-19, the date on which the first such illness began (where they could report any combination of a wide range of symptoms), any previous PCR or antibody test for SARS-CoV-2 (and results), and other details including any illness in their household.

Individuals were subsequently directed to an online booking system for attending a workplace clinic. Availability was given on a first-come-first-served basis, representing a convenience rather than consecutive sample. The online booking system was closed once the pre-specified sample size had been reached. At the clinic appointment, the individual had the opportunity to ask questions, and was asked to sign a re-consent form. A venous blood sample was taken, which was linked back to their questionnaire via a unique study number. Study launch coincided with the launch of the NHS Staff screening programme in England.

### Eligibility criteria

Only individuals who were currently working at their place of work; aged 18 years and over; able to read English (so as to be able to understand the participant information leaflet); and had an in-use personal email address and mobile phone number were eligible to enrol. Anyone who was currently experiencing COVID-19 compatible symptoms; who had experienced such symptoms in the last 7 days; who met the government’s criteria for “exceptionally vulnerable” and as such should be “shielding” (self-isolating); unable to read normal sized print with glasses or; currently taking part in any COVID-19 vaccine trials were not considered eligible for enrolment.

### Serological analysis

A volume of 6mL EDTA anticoagulated blood was taken from each participant and sent to PHE Seroepidemiology Unit (SEU) in Manchester each day for plasma separation and sample banking. Samples were processed by the SEU on the day of receipt by centrifugation at 1200g for 15 minutes. Two approximately 2mL aliquots of plasma were removed and these aliquots stored at –80°C for up to one week. At the end of the week of collection, a single aliquot of each of the samples was shipped in ambient conditions overnight to PHE Porton Down. The second aliquot of each sample was retained at –80°C in the SEU Manchester for future investigations.

Samples were received by PHE Porton Down on 17 June, 19 June, 02 July and 24 July. All samples were analysed using Roche Elecsys ® Anti-SARS-CoV-2 (nucleocapsid (N))^15^ and EUROIMMUN Anti-SARS-CoV-2 ELISA (IgG) assays (Spike (S) protein S1 domain)^17^ Samples were run on EUROIMMUN assay within 24 hours of sample receipt. The samples were then stored at 4°C until there was capacity to run on the Roche assay, which was done between 4 and 16 days following sample receipt.

Manufacturer cut-offs were used to assess serological positivity. A positive result was considered as COI ≥ 1.0 for Roche Elecsys ®^15^. For EUROIMMUN the lower borderline cut-off was used (Ratio > 0.8) rather than upper borderline (Ratio > 1.1)^6^; i.e. any score considered borderline by manufacturer cut-offs was considered a positive within this cohort. Thresholds were pre-specified for both tests. For the purpose of assessing the relationship between clinical signs and symptoms and seropositivity and estimating seroprevalence amongst sub-cohorts, an individual was considered seropositive if they were positive on either assay. Staff conducting the serological analysis did not have access to participant information.

### WHO case definitions as adapted to the UK context

Participants were characterised and reported by an adapted version of the World Health Organization (WHO) criteria for confirmed, suspected and probable cases^18^. This adaptation reflects UK screening and swabbing practices. On 5 March, the UK government confirmed there was evidence of ongoing community transmission; as such, the policy moved from one of “containment” to one of “delay” where testing was initially largely confined to hospitalised cases. This date cut-off was used to characterise cases as follows:

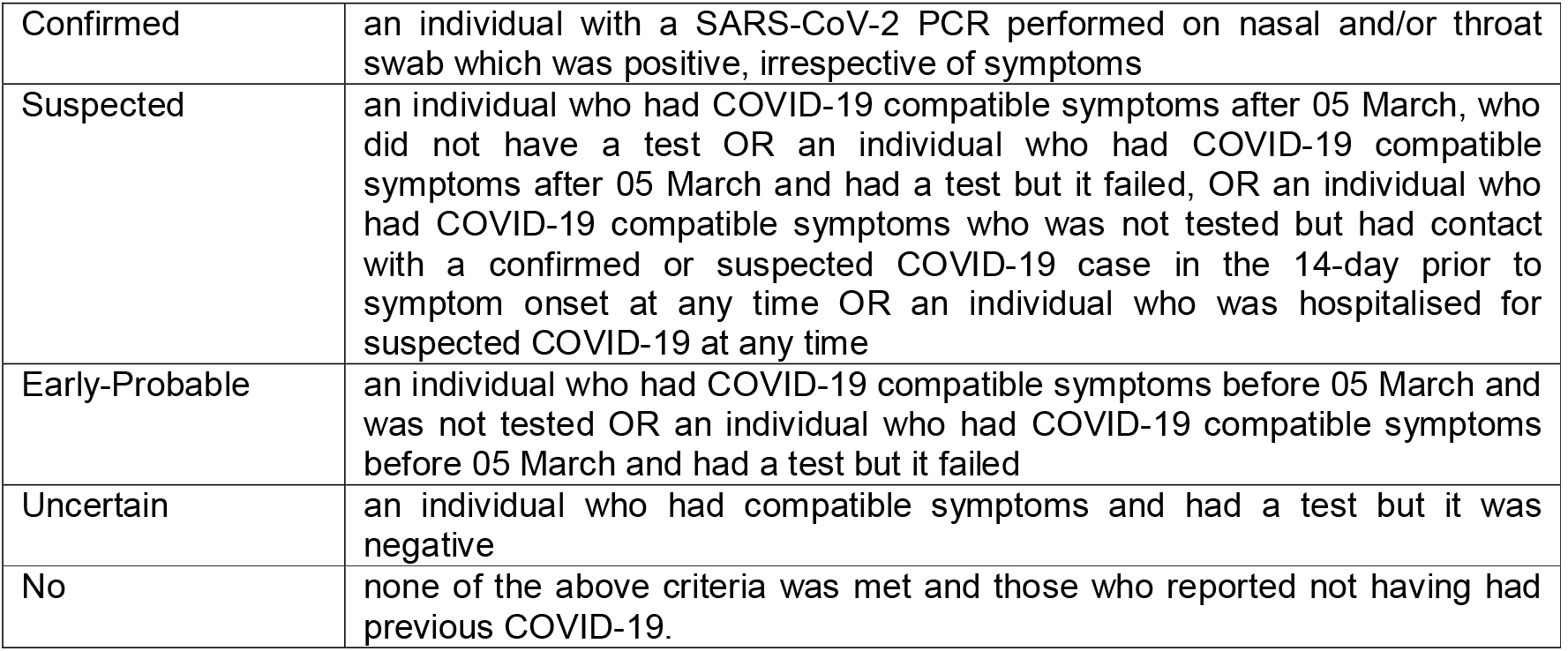

|  |  |
| --- | --- |
| Confirmed | an individual with a SARS-CoV-2 PCR performed on nasal and/or throat swab which was positive, irrespective of symptoms |
| Suspected | an individual who had COVID-19 compatible symptoms after 05 March, who did not have a test OR an individual who had COVID-19 compatible symptoms after 05 March and had a test but it failed, OR an individual who had COVID-19 compatible symptoms who was not tested but had contact with a confirmed or suspected COVID-19 case in the 14-day prior to symptom onset at any time OR an individual who was hospitalised for suspected COVID-19 at any time |
| Early-Probable | an individual who had COVID-19 compatible symptoms before 05 March and was not tested OR an individual who had COVID-19 compatible symptoms before 05 March and had a test but it failed |
| Uncertain | an individual who had compatible symptoms and had a test but it was negative |
| No | none of the above criteria was met and those who reported not having had previous COVID-19. |

These definitions were pre-specified; further details are in Supplementary Material.

### Statistical analysis

All data were collected via an online questionnaire (Snap Survey), and were stored and managed on a secure server using a relational database management system (SQL). Data were cleaned and analysed using R version 3.5.1. Descriptive analysis was conducted to describe the cohort by age, gender, ethnicity, occupation, recent symptoms compatible with COVID-19 and previous known exposure to SARS-CoV-2 and by results on the two anti-SARS-CoV-2 immunoassays. Comparison was made with laboratory confirmed COVID-19 data for England obtained from http://data.coronavirus.gov.uk on 28 July 2020.

To understand the relationship between reported symptoms, symptoms were coded as 1 (present) or 0 (not present), and a matrix of Pearson correlation coefficients computed. This reflects correlations between personal symptoms and those in household contacts. This matrix was depicted using the R corrplot package, following hierarchical clustering of the matrix (R hclust function, with Euclidean distance metric and central clustering^19^).

Univariable logistic regression was conducted to determine unadjusted odds ratios (ORs) with 95% confidence intervals of seropositivity given various clinical risk factors and seropositivity for those in Police and Fire and HCW groups.

Sensitivity of the immunoassays was estimated by the proportion of confirmed cases (individuals who had a previously PCR-positive nasal or throat swab) with detectable SARS-CoV-2 antibodies. This is reported for all cases, and by categories of time of testing since symptom onset. As antibody assay signals are not normally distributed, when comparing antibody assay signals non-parametric tests were used. Spearman’s rank correlation coefficient was used to assess the concordance between the two assays. Quantitative scores for the two assays were also plotted against time between date of symptom onset and the date when the blood sample was taken for HCW-PP (illness-venesection interval). Quantile regression (R quantreg package) was used to test for evidence for a decline over time. To assess the relationship between duration of symptoms (a categorical variable) and antibody assay signal (a continuous variable) we used Kendall’s correlation coefficient.

### Patient and public involvement statement

Contributors from patient and public involvement groups at NIHR Birmingham Biomedical Research Centre and NIHR Applied Research Collaboration West Midlands were involved in reviewing the proposed study documents through the HRA fast track review process set up for urgent COVID-19 studies.

## Results

### Recruitment

Recruitment opened on 27 May 2020, and study clinics ran from 1 June to 26 June 2020 in two nonhealthcare worker sites (one Police and one Fire & Rescue) and in six NHS acute hospitals (Supplementary Figure 1). 3,087 individuals completed a questionnaire, of whom 2867 (93%) attended a clinic appointment and were successfully enrolled into the study. We excluded individuals due to non-eligibility (n = 1, who was recruited in “Stream C” however did not have a previously positive PCR result), technical issues (e.g. insufficient sample) preventing blood samples being analysed (n = 14), and withdrawals from the study after recruitment (n = 5). There were no test failures on either immunoassay. The final cohort contained 2,847 individuals: 1,147 from Police and Fire (Stream A); 1,546 health care workers (HCW) (Stream B); and 154 from the healthcare worker previously COVID-19 positive test group (HCW-PP) (Stream C) (Figure 1).

**Figure 1:**
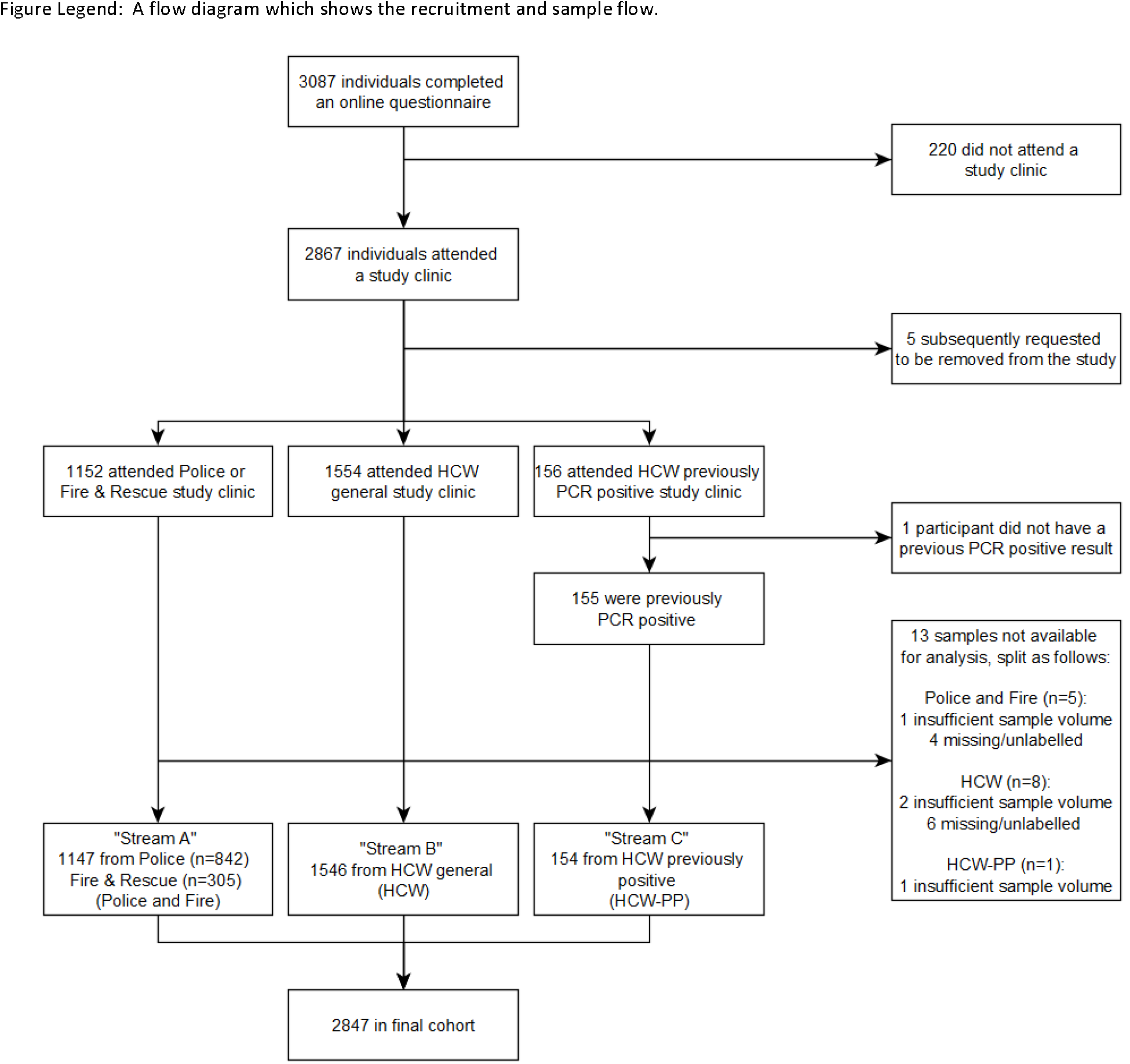
Recruitment and sample analysis flow diagram

### Cohort description

Of the 2,847 individuals, 36% were male and 64% were female. Their ages ranged from 19 to 73 years with a median age of 43 (Supplementary Figure 2). Ethnicity was majority white (83%), followed by 10% Asian, 3% Black, 2% Mixed and 2% Other. Overall 44% reported face to face interactions during lockdown with clients/patients at a similar frequency to pre-lockdown. At the time the questionnaire was administered, only 3% had had a COVID-19 antibody test and had been informed of the result. All the HCW-PP group had (by definition) a previously positive SARS-CoV-2 PCR result; by contrast, only 2% of the Police and Fire group and 6% of the HCW group had a prior PCR positive result.

**Figure 2:**
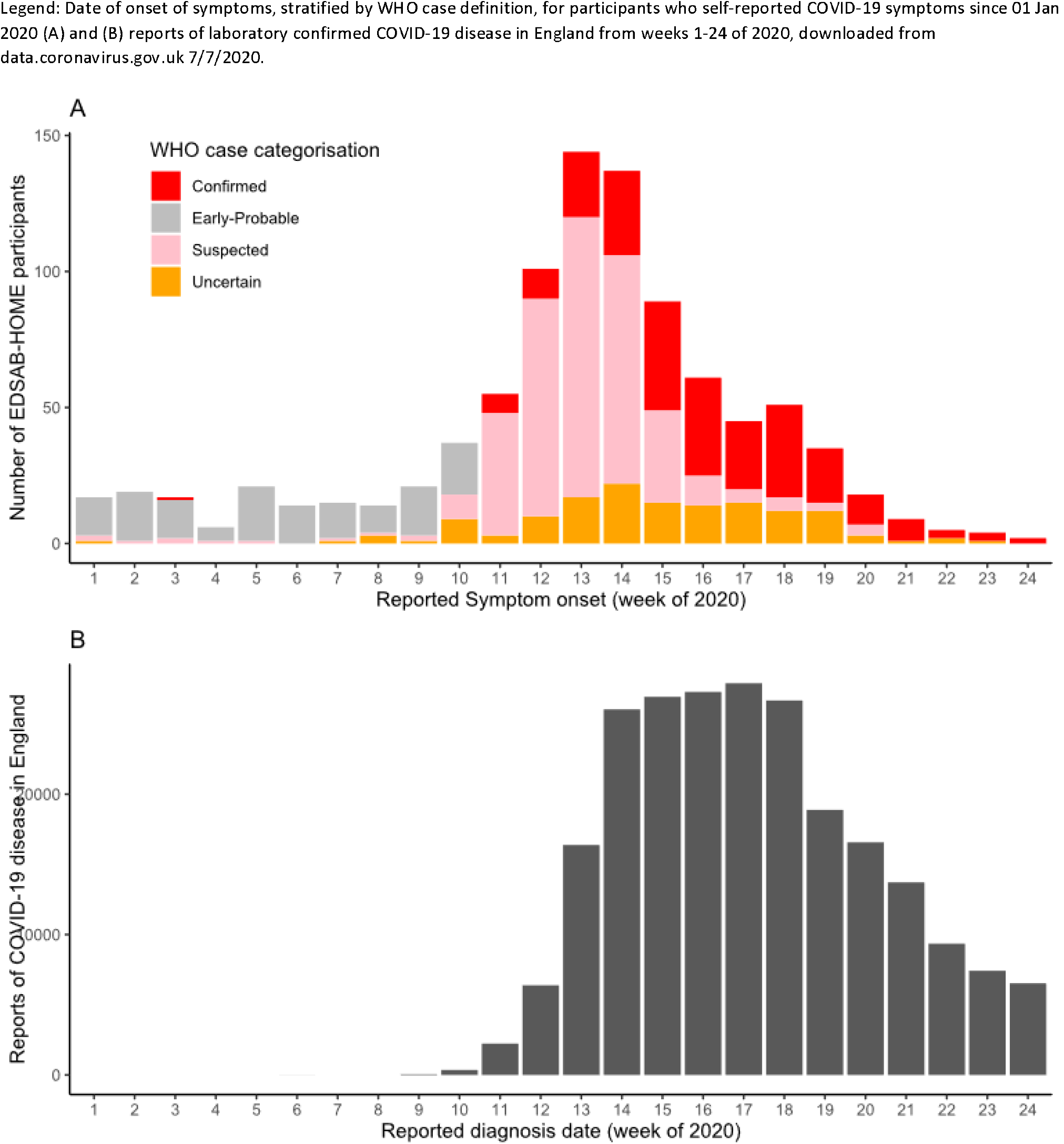
Epidemiological curve for participants reporting symptoms

Overall, 943 (33%) participants reported that they believed they had already had COVID-19 due to compatible symptoms; this belief was present in the HCW-PP, Police and Fire and HCW groups in 99% (the remaining 1% were asymptomatic), 26% and 32% respectively (Table 1). Among individuals who reported symptoms, the median time since symptoms onset to the EDSAB-HOME recruitment visit was 75 days (IQR 63 – 92 days), which was on average 12 days longer than the time since symptoms among the known previous PCR positives (median 63 days). In this cohort, hospitalisation due to COVID-19 like illness was unusual, being reported in 1% and 4% of Fire & Police and HCW groups, respectively (Table 1).

**Table 1:**
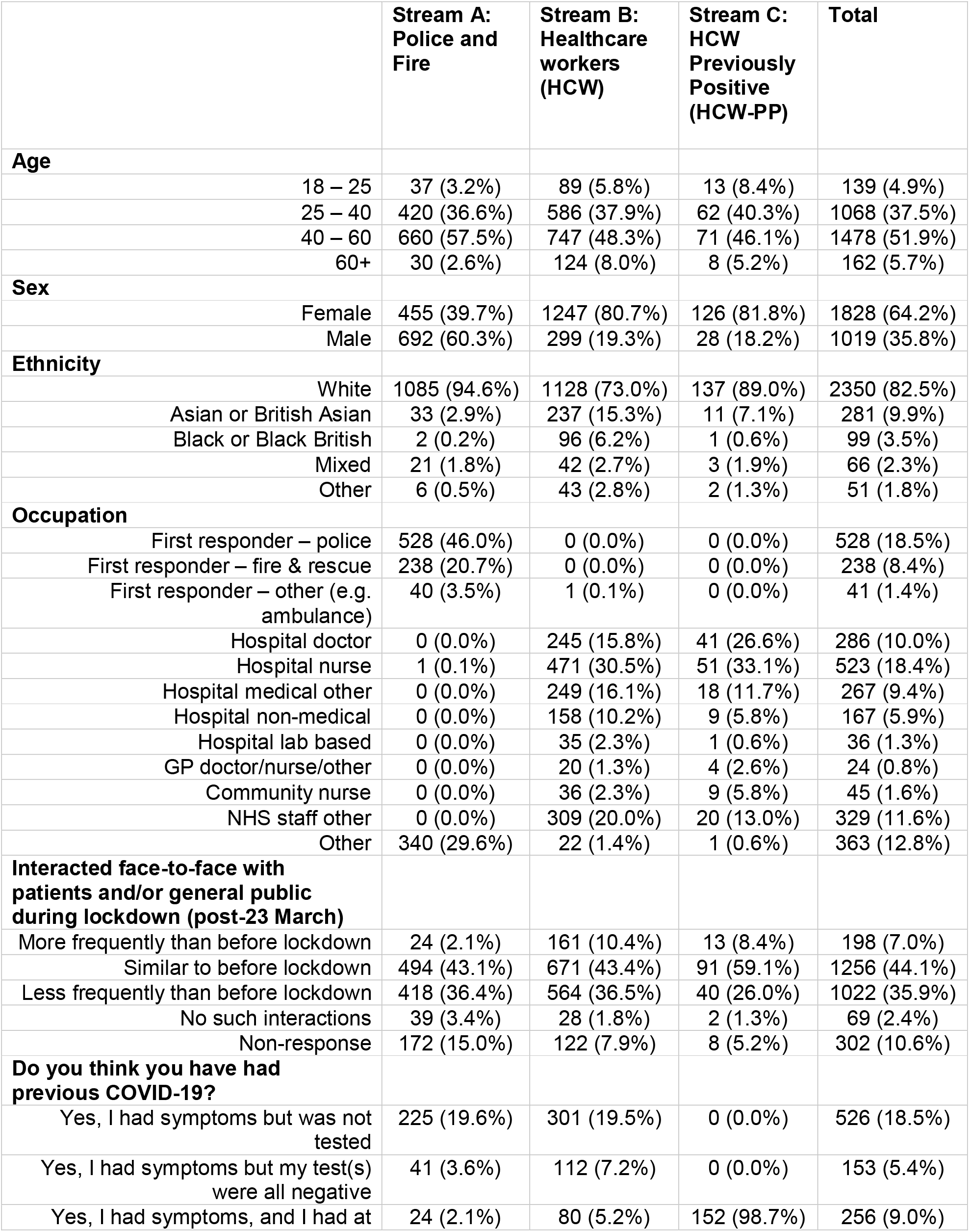
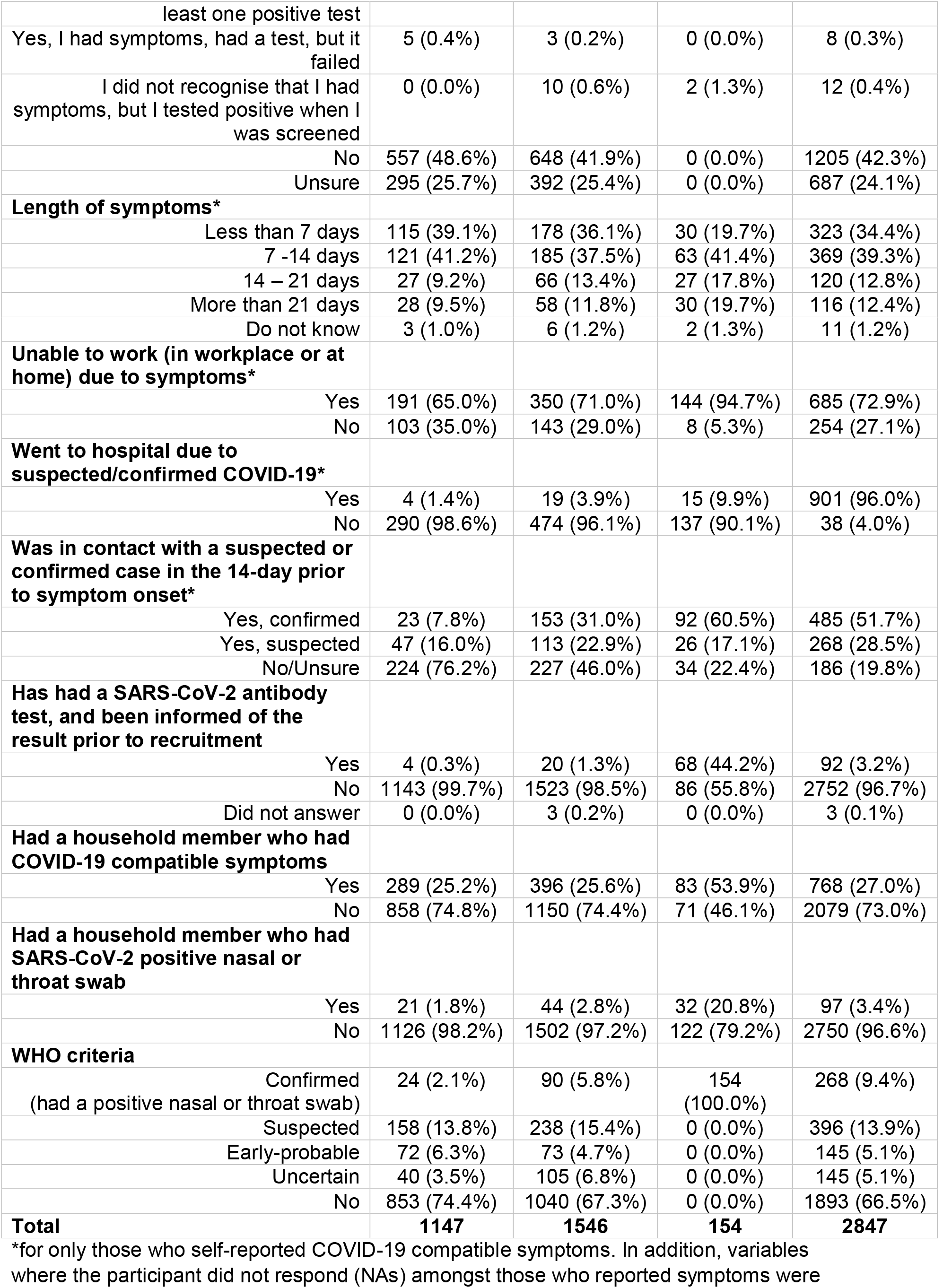
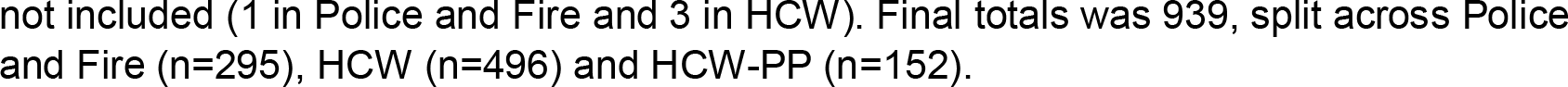
Demographics and exposure characteristics, split by recruitment group.

Of all participants, 9% were in the WHO “confirmed” category, 14% were “suspected”, 5% were “early-probable” cases, and 5% were categorised as “uncertain”. Of the “confirmed” cases, only 12 were identified through screening (2 in HCW-PP and 10 in HCW). Reported onset dates are shown in Figure 2.

### Symptom correlations

Some symptoms were commonly reported together (Figure 3). For example, altered sense of taste (ageusia) and smell (anosmia) were commonly co-reported; a symptom cluster of breathlessness, cough, muscle aching, fever and fatigue was also evident. Reporting symptoms in both these clusters was strongly associated with both belief that one had had COVID-19, and with seropositivity. Much weaker associations with seropositivity and with belief in prior COVID-19 infection were observed with other symptom clusters identified, and with clusters of symptoms reported in household members.

**Figure 3:**
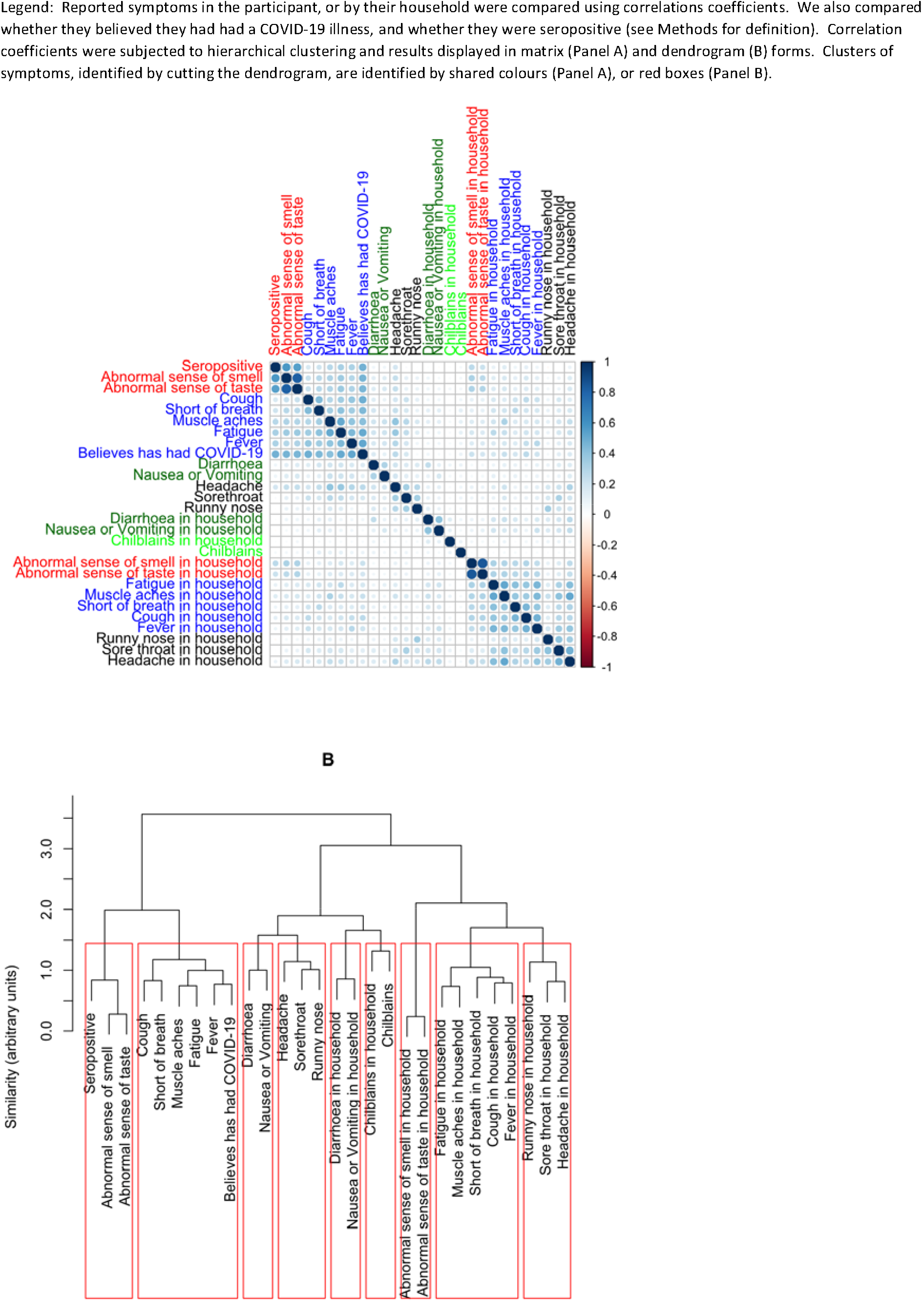
Symptoms reported by participants

### Seropositivity and its relationship to belief in past COVID-19 infection

Seropositivity was more common amongst those who reported having had COVID-19 compatible symptoms: amongst those who self-reported COVID-19 compatible symptoms, 28% in Police and Fire and 50% of healthcare workers were seropositive compared with 5% and 11% of those who did not report such symptoms (Table 2). Therefore, self-belief of having had COVID-19 was associated with seropositivity, in both the Police and Fire (OR 7.5 95% CI 5.0–11.3) and in HCW (OR 7.9 95% 6.1–10.3). However, of those seropositive across both groups, only 68% thought they had had COVID-19 (equating to 32% of the seropositives having had asymptomatic COVID-19).

**Table 2:**
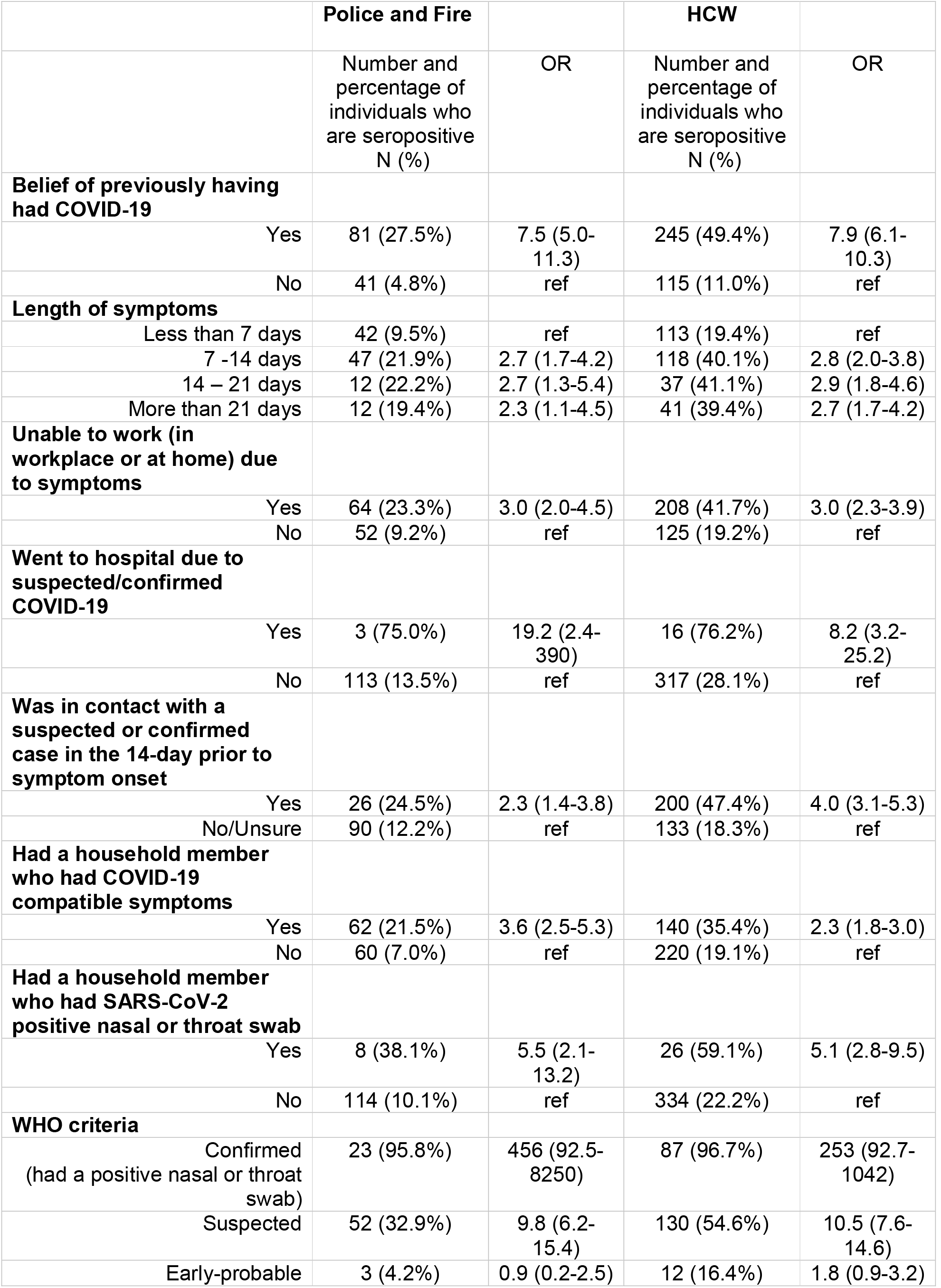
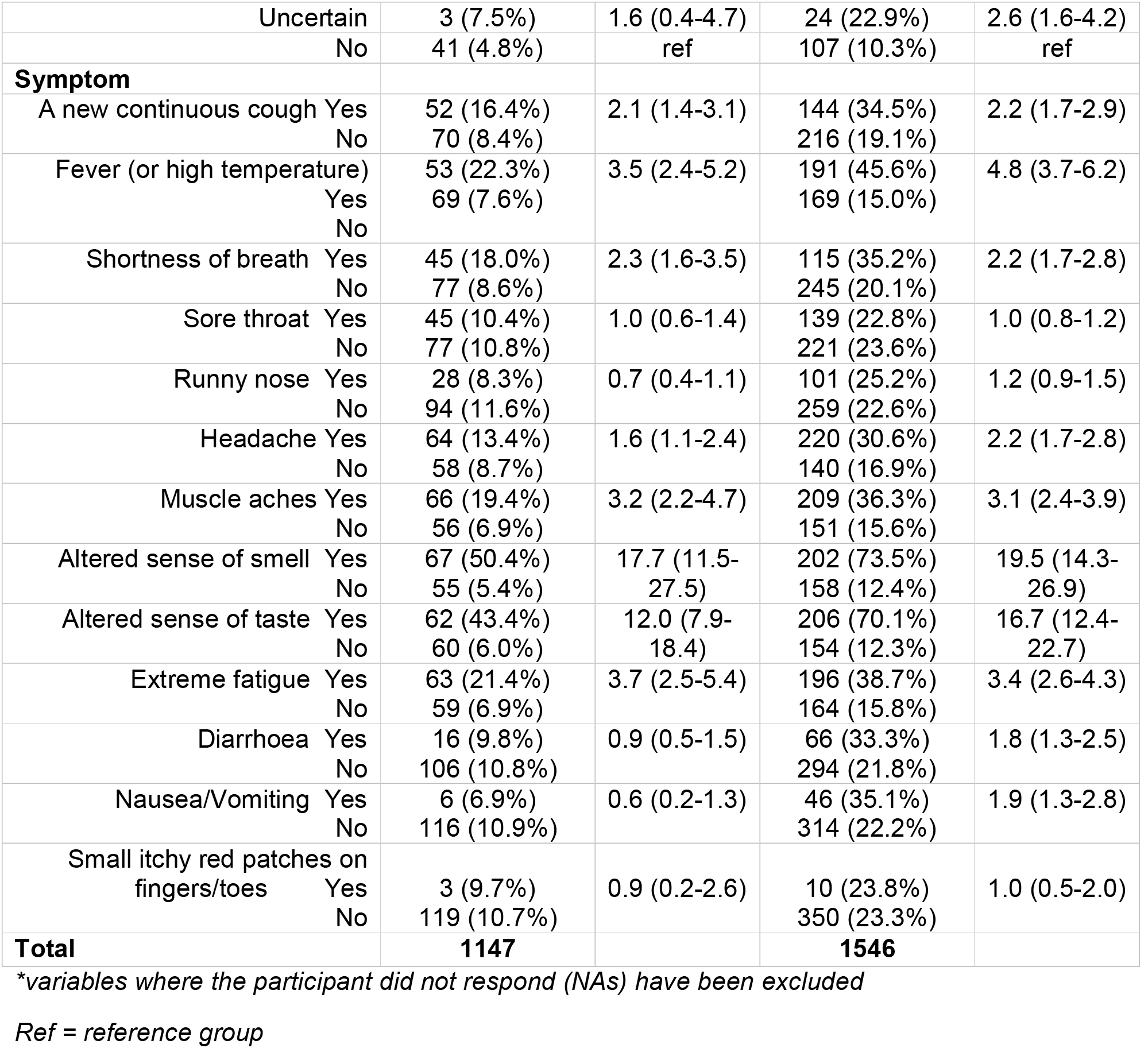
Seropositivity by recruitment group for clinical risk factors*.

|  | Police and Fire |  | HCW |  |
| --- | --- | --- | --- | --- |
|  | Number and percentage of individuals who are seropositive<br>N (%) | OR | Number and percentage of individuals who are seropositive<br>N (%) | OR |
| **Belief of previously having had COVID-19** |  |  |  |  |
| Yes | 81 (27.5%) | 7.5 (5.0-11.3) | 245 (49.4%) | 7.9 (6.1-10.3) |
| No | 41 (4.8%) | ref | 115 (11.0%) | ref |
| **Length of symptoms** |  |  |  |  |
| Less than 7 days | 42 (9.5%) | ref | 113 (19.4%) | ref |
| 7 -14 days | 47 (21.9%) | 2.7 (1.7-4.2) | 118 (40.1%) | 2.8 (2.0-3.8) |
| 14 – 21 days | 12 (22.2%) | 2.7 (1.3-5.4) | 37 (41.1%) | 2.9 (1.8-4.6) |
| More than 21 days | 12 (19.4%) | 2.3 (1.1-4.5) | 41 (39.4%) | 2.7 (1.7-4.2) |
| **Unable to work (in workplace or at home) due to symptoms** |  |  |  |  |
| Yes | 64 (23.3%) | 3.0 (2.0-4.5) | 208 (41.7%) | 3.0 (2.3-3.9) |
| No | 52 (9.2%) | ref | 125 (19.2%) | ref |
| **Went to hospital due to suspected/confirmed COVID-19** |  |  |  |  |
| Yes | 3 (75.0%) | 19.2 (2.4-390) | 16 (76.2%) | 8.2 (3.2-25.2) |
| No | 113 (13.5%) | ref | 317 (28.1%) | ref |
| **Was in contact with a suspected or confirmed case in the 14-day prior to symptom onset** |  |  |  |  |
| Yes | 26 (24.5%) | 2.3 (1.4-3.8) | 200 (47.4%) | 4.0 (3.1-5.3) |
| No/Unsure | 90 (12.2%) | ref | 133 (18.3%) | ref |
| **Had a household member who had COVID-19 compatible symptoms** |  |  |  |  |
| Yes | 62 (21.5%) | 3.6 (2.5-5.3) | 140 (35.4%) | 2.3 (1.8-3.0) |
| No | 60 (7.0%) | ref | 220 (19.1%) | ref |
| **Had a household member who had SARS-CoV-2 positive nasal or throat swab** |  |  |  |  |
| Yes | 8 (38.1%) | 5.5 (2.1-13.2) | 26 (59.1%) | 5.1 (2.8-9.5) |
| No | 114 (10.1%) | ref | 334 (22.2%) | ref |
| **WHO criteria** |  |  |  |  |
| Confirmed<br>(had a positive nasal or throat swab) | 23 (95.8%) | 456 (92.5-8250) | 87 (96.7%) | 253 (92.7-1042) |
| Suspected | 52 (32.9%) | 9.8 (6.2-15.4) | 130 (54.6%) | 10.5 (7.6-14.6) |
| Early-probable | 3 (4.2%) | 0.9 (0.2-2.5) | 12 (16.4%) | 1.8 (0.9-3.2) |

|  |  |  |  |  |  |
| --- | --- | --- | --- | --- | --- |
| Uncertain |  | 3 (7.5%) | 1.6 (0.4-4.7) | 24 (22.9%) | 2.6 (1.6-4.2) |
| No |  | 41 (4.8%) | ref | 107 (10.3%) | ref |
| **Symptom** |  |  |  |  |  |
| A new continuous cough | Yes | 52 (16.4%) | 2.1 (1.4-3.1) | 144 (34.5%) | 2.2 (1.7-2.9) |
|  | No | 70 (8.4%) |  | 216 (19.1%) |  |
| Fever (or high temperature) | Yes | 53 (22.3%) | 3.5 (2.4-5.2) | 191 (45.6%) | 4.8 (3.7-6.2) |
|  | No | 69 (7.6%) |  | 169 (15.0%) |  |
| Shortness of breath | Yes | 45 (18.0%) | 2.3 (1.6-3.5) | 115 (35.2%) | 2.2 (1.7-2.8) |
|  | No | 77 (8.6%) |  | 245 (20.1%) |  |
| Sore throat | Yes | 45 (10.4%) | 1.0 (0.6-1.4) | 139 (22.8%) | 1.0 (0.8-1.2) |
|  | No | 77 (10.8%) |  | 221 (23.6%) |  |
| Runny nose | Yes | 28 (8.3%) | 0.7 (0.4-1.1) | 101 (25.2%) | 1.2 (0.9-1.5) |
|  | No | 94 (11.6%) |  | 259 (22.6%) |  |
| Headache | Yes | 64 (13.4%) | 1.6 (1.1-2.4) | 220 (30.6%) | 2.2 (1.7-2.8) |
|  | No | 58 (8.7%) |  | 140 (16.9%) |  |
| Muscle aches | Yes | 66 (19.4%) | 3.2 (2.2-4.7) | 209 (36.3%) | 3.1 (2.4-3.9) |
|  | No | 56 (6.9%) |  | 151 (15.6%) |  |
| Altered sense of smell | Yes | 67 (50.4%) | 17.7 (11.5-27.5) | 202 (73.5%) | 19.5 (14.3-26.9) |
|  | No | 55 (5.4%) |  | 158 (12.4%) |  |
| Altered sense of taste | Yes | 62 (43.4%) | 12.0 (7.9-18.4) | 206 (70.1%) | 16.7 (12.4-22.7) |
|  | No | 60 (6.0%) |  | 154 (12.3%) |  |
| Extreme fatigue | Yes | 63 (21.4%) | 3.7 (2.5-5.4) | 196 (38.7%) | 3.4 (2.6-4.3) |
|  | No | 59 (6.9%) |  | 164 (15.8%) |  |
| Diarrhoea | Yes | 16 (9.8%) | 0.9 (0.5-1.5) | 66 (33.3%) | 1.8 (1.3-2.5) |
|  | No | 106 (10.8%) |  | 294 (21.8%) |  |
| Nausea/Vomiting | Yes | 6 (6.9%) | 0.6 (0.2-1.3) | 46 (35.1%) | 1.9 (1.3-2.8) |
|  | No | 116 (10.9%) |  | 314 (22.2%) |  |
| Small itchy red patches on fingers/toes | Yes | 3 (9.7%) | 0.9 (0.2-2.6) | 10 (23.8%) | 1.0 (0.5-2.0) |
|  | No | 119 (10.7%) |  | 350 (23.3%) |  |
| **Total** |  | **1147** |  | **1546** |  |

*\*variables where the participant did not respond (NAs) have been excluded*
*Ref = reference group*

### Seropositivity and reported symptoms

Our study detected seropositivity as associated with several known clinical risk factors (Table 2). In both Police and Fire and HCW, having symptoms longer than 7 days, being unable to work due to symptoms, previous hospitalisation, having had a household member who had COVID-19 compatible symptoms, having had a household member who had a positive nasal or throat swab, and having symptoms of cough, fever, shortness of breath, headache, muscle aches, altered sense of smell or taste and extreme fatigue were all associated with seropositivity (Table 2, Figure 3, Supplementary Figure 3). The association varied for individual symptoms and seropositivity, such as altered sense of smell (OR 19.5 (95%CI 14.3–26.9) and taste (OR 16.7 (95%CI 12.4–22.7), and new continuous cough (OR 2.2 (95%CI 1.7–2.9)).

We also noted that date of symptom onset was associated with likelihood of seropositivity: while 46% of the “suspected” (symptoms after 5 March) group were seropositive, only 10% of “early-probable” (symptoms up to 5 March, which predate the national outbreak) were seropositive (Figure S3). Of the “uncertain” group, 19% were seropositive, while 8% were seropositive in those reporting no symptoms.

Overall, amongst those who reported a belief of having had COVID-19, a higher proportion of those in Police and Fire and HCW reported symptoms of fewer than 7 days (39%, 36%) than those in the HCW-PP group with microbiologically confirmed COVID-19 (20%). A lower proportion were unable to work due to their symptoms (65% in Police and Fire and 71% HCW vs 95% HCW-PP) or were hospitalised (1%, 4% vs 10% respectively) (as such assumed to have more mild symptoms). A lower proportion had been in contact with a confirmed case in the 14-days prior to symptom onset (8% and 31% vs 60% respectively) or had a household member with COVID-19 compatible symptoms (25% and 26% vs 53% respectively) (Table 1). Thus, we observed that those reporting symptoms who were seronegative differed in multiple respects from those who reported symptoms and were seropositive.

Amongst individuals not reporting any previous COVID-19 compatible symptoms (or any previous PCR test), 158 (8%) were seropositive. Amongst these, 81% reported that no household member (or did not know) had any COVID-19 compatible symptoms; together, these data highlight the potential of serology to identify (albeit to an unknown degree), previously unidentified asymptomatic SARS-CoV-2 infection.

### Serological assay performance

One explanation for the large numbers of symptomatic individuals with negative serology might be the sensitivity of the assays^7^. In our study, anti-Spike (EUROIMMUN) (Anti-S1) indices and Anti-Nucleocapsid (Roche) (Anti-N) protein assay ratios showed a strong positive correlation (r = 0.93, p < 0.001) and each had a clear bimodal distribution within streams A and B (Figure 4).

**Figure 4:**
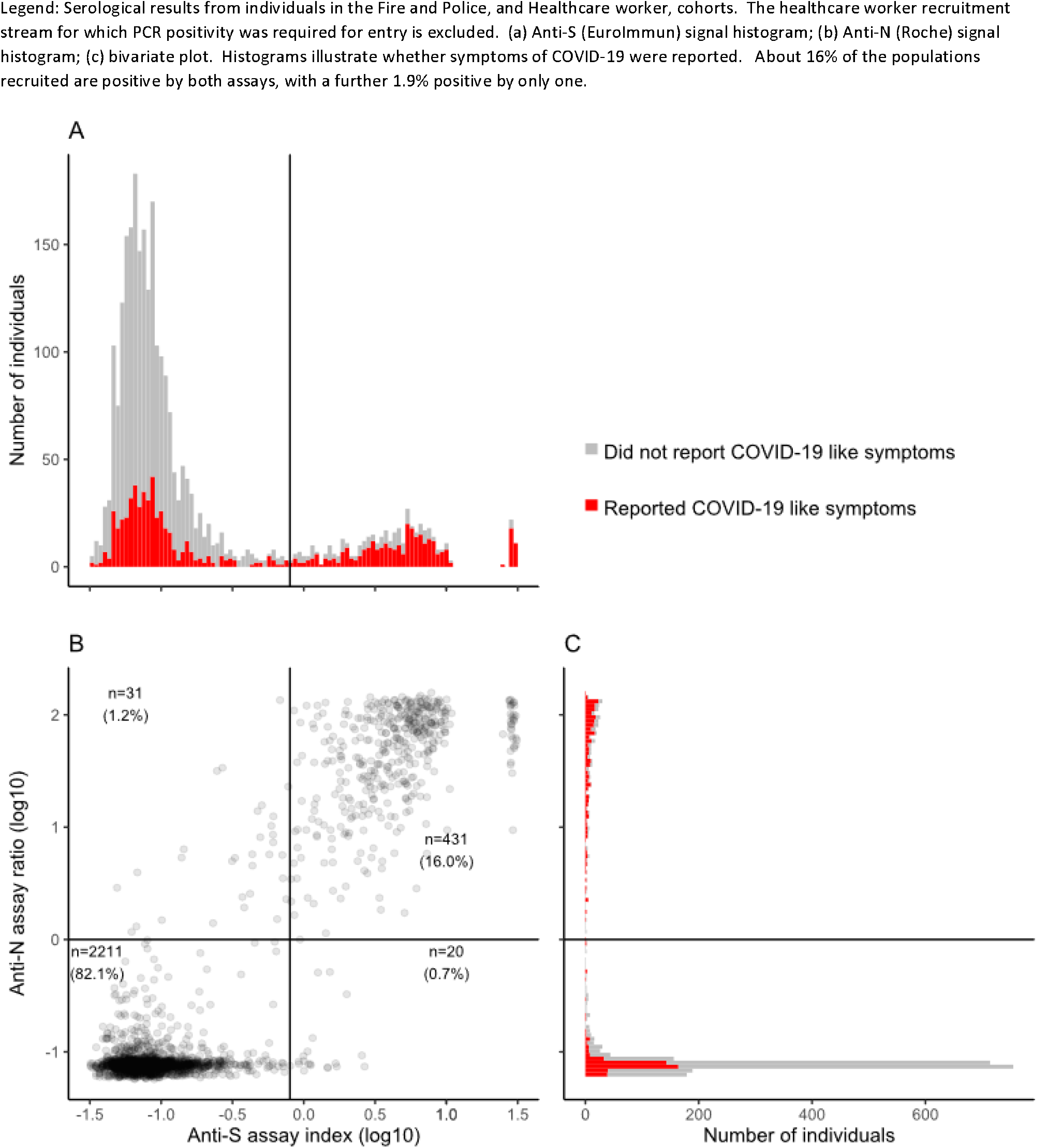
Serological results in individuals from Fire and Police and Healthcare worker cohorts.

There were in total 268 individuals who had previous PCR test positive infection (all 154 from HCWPP C, plus 24 from Police and Fire and 90 from HCW) (Figure 5, Supplementary Table 1). The majority were white (n = 212, 79%) and female (n = 188, 70%). 4% (n = 12) had been identified through screening and were asymptomatic at time of swab. 11% had been hospitalised, and all were on average 63 days (IQR 52 – 75 days) post symptom onset. 65% had illness lasting 2 weeks or less. Based on these 268 individuals, the sensitivity of the anti-S1 and anti-N assays was 93.3% (95% CI 89.6% – 95.7%) and 96.6% (95% CI 93.7% – 98.2%), respectively. The composite sensitivity (i.e. proportion of positive on at least one assay) was 98.1% (95% CI 95.7% – 99.2%). Of the 12 individuals who had been asymptomatic when they had their positive PCR test, 8 were seropositive across both assays (Of the remaining 4, 1 was positive on EUROIMMUN only, while 3 were negative across both).

**Figure 5:**
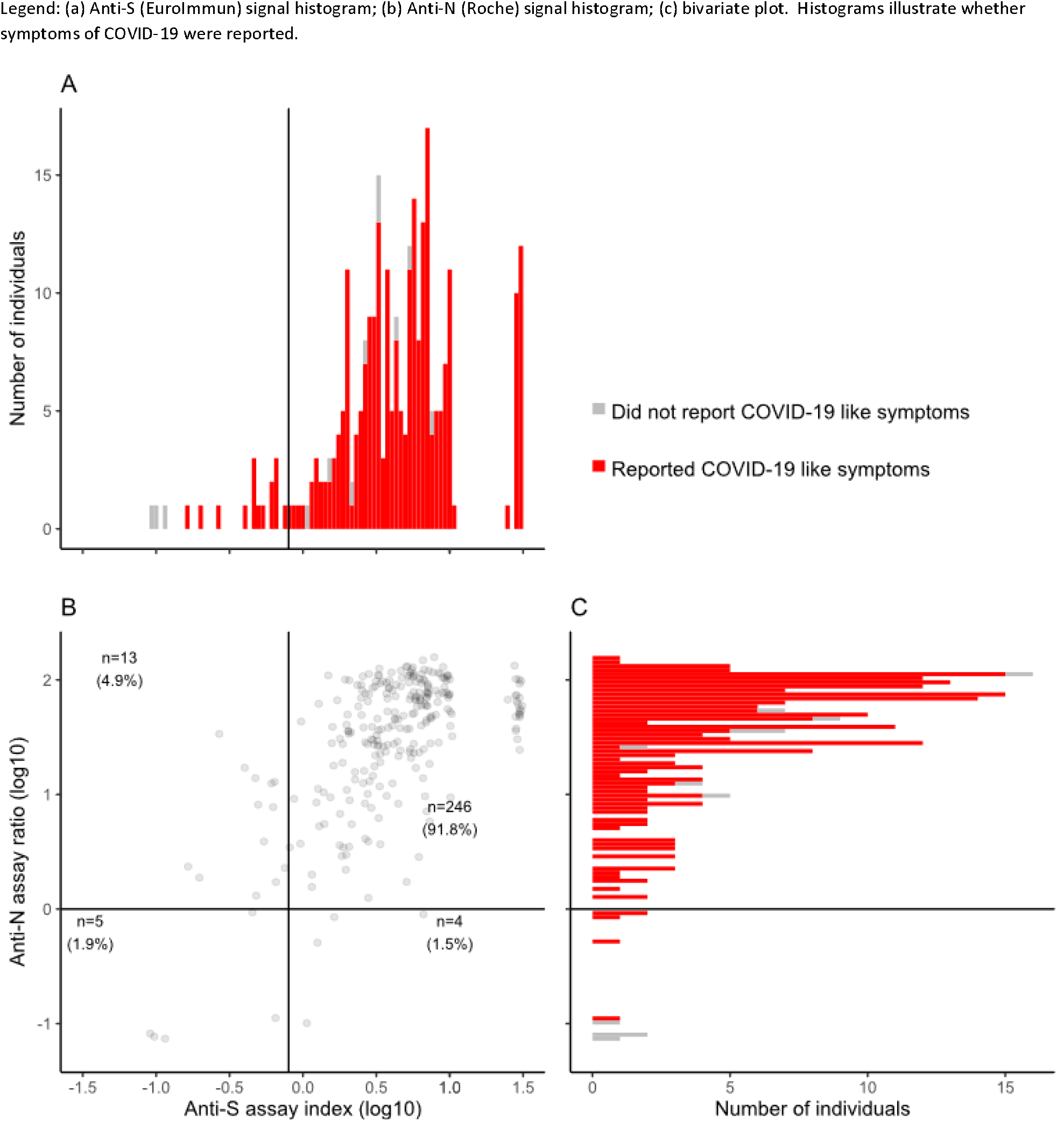
Serological results in individuals with previous PCR positivity (confirmed cases).

### Do antibody assay signals decline?

There is weak statistical evidence for higher anti-S antibody signals in individuals with longer illness duration (*p* = 0.06; for anti-N, *p* = 0.16, Figure 6A, C). There is also evidence for a decline over time for anti-S antibody signals with symptom onset to blood sample intervals of between 35 and 110 days, intervals chosen as they include 87% of symptomatic known previous PCR positive cases (n = 233). Regression models estimate a decline per month of 29.1% (95% CI 3.1% to 36.7%) (Figure 6B), with similar results in models adjusting for initial illness duration (not shown). There was a similar trend in anti-N signals (with an estimated 18.7% decline per month, 95% CI –26.2% to 36.9%), however this was not statistically significant (Figure 6D). Over the time period studied, this decline has minimal impact on the serological detection of individuals with microbiologically confirmed COVID-19 disease (Supplementary Table 2). In summary, neither serological assay sensitivity, nor declining antibody titres, appear to provide an explanation for approximately half of symptomatic individuals who are antibody negative in our cohort.

**Figure 6:**
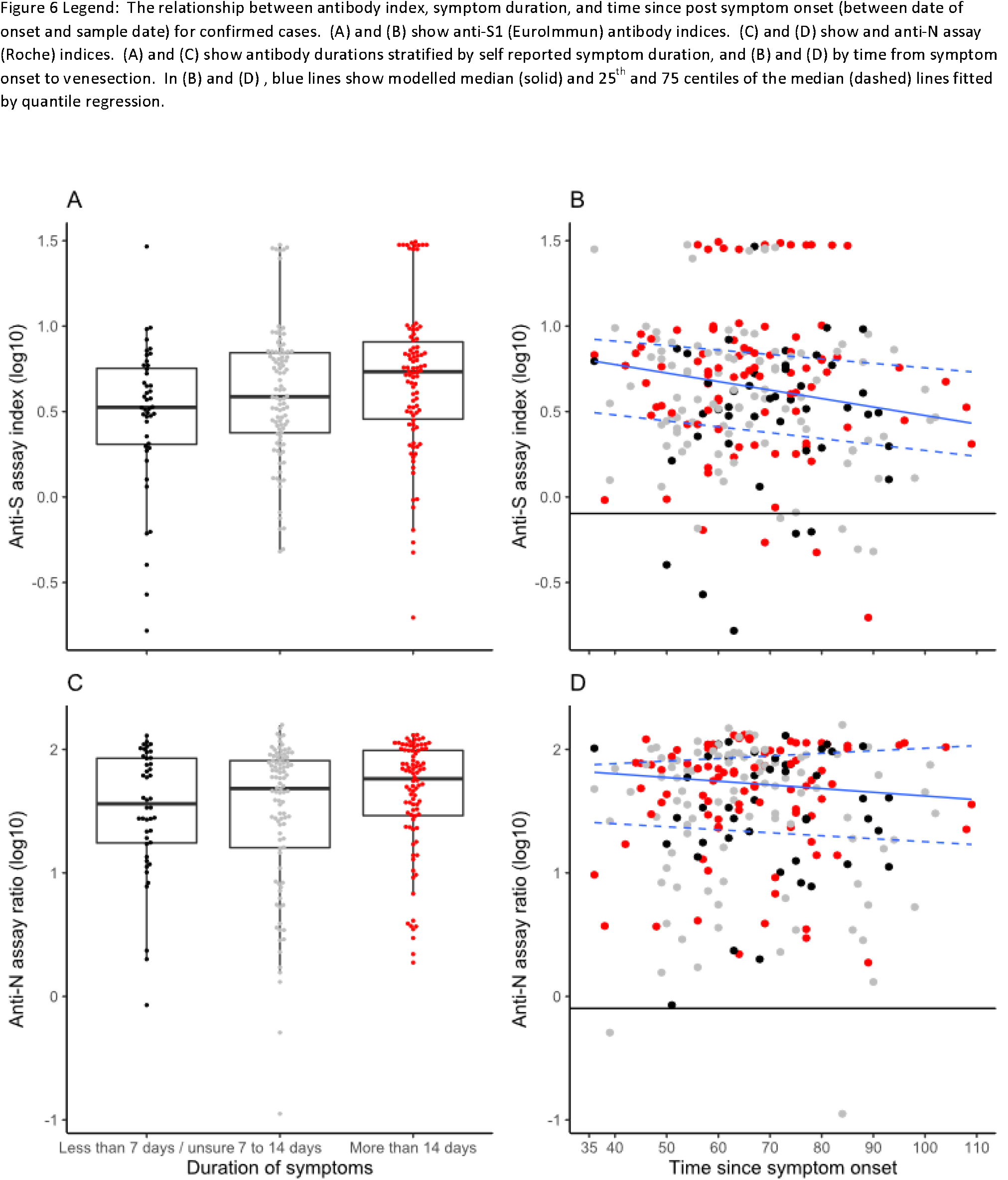
Antibody responses, illness duration and interval post illness

## Discussion

In summary, in our study of key workers, performed concomitant with the start of the NHS staff antibody testing programme, we observed that a high proportion (∼33%) believe they had had COVID-19 due to compatible symptoms, but that, of these, around half lacked any serological evidence of having had the infection. Amongst individuals who had had symptoms, those who were seronegative reported earlier dates of symptom onset, shorter duration of symptoms and were less likely to report high specificity COVID-19 symptoms of ageusia and anosmia.

The high proportion of individuals reporting belief they had had COVID-19 but who were seronegative did not appear to be readily explicable by low serological assay sensitivity or rapidly declining antibody titres. We estimated that both immunoassays used had high sensitivity (93.3%, CI 89.6% –95.7%; and 96.7%, CI 93.7% – 98.2%, for EUROIMMUN and Roche assays, respectively) in the frontline worker populations studied, who were on average 63 days post PCR-confirmed infection. This is despite many of these individuals having had relatively mild disease: among the 268 previously PCR positive individuals studied, only about 10% had been hospitalised.

A strength of our study is the detailed symptom history provided by a large cohort of keyworker participants. Although there is the potential for recall bias, about 97% of the cohort provided symptom data naïve to their serological status, and it would appear (perhaps due to interest in COVID-19 during the ongoing pandemic) that retrospective symptom collection has yielded accurate data: symptom onsets reported mirror the UK outbreak among seropositive (but not seronegative) cases, and symptom combinations reported appear to be strongly predictive of COVID-19, such as the now well-known combinations of altered taste and smell^10^. In addition, several other self-reported factors which are known to increase likelihood of infection, such as having had contact with a confirmed/suspected COVID-19 case and having a household member who had a positive swab were associated with seropositivity.

Comparing the data obtained with published data, the test sensitivity estimates we derive are compatible with published results for individuals with mild disease (and who had met the threshold for testing)^20 5^, but are higher than those reported by laboratory assessments using smaller panels with few individuals > 21 days post infection^15,17^, likely due to the increased time since symptom onset (median 63 days in this cohort) relative to that in previous evaluations. This extended internal has allowed, both antibody concentrations, and perhaps also affinity of antibodies for SARS-CoV-2^21,22^ to rise. Our results are also congruent with reports from anti-Spike S1 Receptor binding domain assays^23^, and with neutralisation assays^16^, in that we observed anti-S titres decline over time post infection. We did not observe significant declines in anti-N immunoassay signals over the time interval studied, but cannot exclude this happening; nevertheless, we can extend the existing literature by showing that these declines have minimal impact on assay performance up to 110 days post infection in symptomatic key workers.

We caution against generalisation to antibody kinetics in other populations, however: the front line key workers we studied, all of whom worked during the pandemic, may have been re-exposed to SARS-CoV-2 during their work, and may have been serially boosted by re-exposure to virus leading to sustained antibody responses, as noted with other viral pathogens^24^.

In terms of implications for police and practice, this study indicates that a substantial proportion of frontline workers believe that they have had COVID-19 based on symptoms experience, but have no antibodies detectable a median of 63 days since symptom onset. As screening programmes are extended across key worker populations, increasing numbers of individuals may receive such unexpected results. Given the differing symptom profiles observed between seropositive and seronegative individuals studied, and the high likelihood of seroconversion in symptomatic illness with confirmed viral presence, it is likely that the unexpected seronegativity is mostly due to a lack of both past infection and any protection such infection might prove to confer. This is something which prospective studies will be able to monitor, allowing determination of whether all protection (if any) from COVID-19 disease can be captured by immunoassays.

## Data Availability

All data is stored securely on a secure PHE server. Data is available on reasonable request via PHE's Office of Data Release (ODR).

## Ethical statement

The study was approved by NHS Research Ethics Committee (Health Research Authority, IRAS 284980) on 02/06/2020 and PHE Research Ethics and Governance Group (REGG, NR0198) on 21/05/2020.

## Acknowledgements

The authors would like to acknowledge key collaborators: Chief Inspector Julie Rawsthorne from National Police Wellbeing Service and Simon Fryer, Area Manager, Lancashire Fire & Rescue Service. In addition, we would like to specifically thank Megan Lamb and Melissa Murove who were integral in ensuring successful recruitment. Finally, we would like to thank all our colleagues from Office of Life Sciences and Department of Health and Social Care who are currently working on providing antibody testing programmes for the UK population. DW acknowledges support from the NIHR Health Protection Research Unit in Genomics and Data Enabling. HEJ, AEA, MH and IO acknowledge support of the NIHR Health Protection Research Unit in Behavioural Science and Evaluation at University of Bristol. STP is supported by an NIHR Career Development Fellowship (CDF-2016–09–018). The views expressed are those of the author(s) and not necessarily those of the NHS, the NIHR or the Department of Health and Social Care.

## Author contributions

Study design – RM, STP, HEJ, AEA, TB, AC, MH, IO, DW

Data collection – RM, DW, PDK

Study site coordination – RS, PM, JB, AH, NT, AC, IR

Processed and analysed immunoassay results – RB, EL, TB

Statistical analysis and expertise – RM, DW, STP, HEJ, TA

Drafted manuscript – RM, DW

Data interpretation – RM, STP, HEJ, AEA, MH, IO, DW

All authors critically reviewed the final paper and agree to be accountable for all aspects of the work and approved the final version for publication.

## Transparency declaration

The lead author affirms that the manuscript is an honest, accurate, and transparent account of the study being reported; that no important aspects of the study have been omitted; and that any discrepancies from the study as planned have been explained.

